# Social Contact Patterns and Implications for Infectious Disease Transmission: A Systematic Review and Meta-Analysis of Contact Surveys

**DOI:** 10.1101/2021.06.10.21258720

**Authors:** Andria Mousa, Peter Winskill, Oliver J Watson, Oliver Ratmann, Mélodie Monod, Marco Ajelli, Aldiouma Diallo, Peter J Dodd, Carlos G Grijalva, Moses Chapa Kiti, Anand Krishnan, Rakesh Kumar, Supriya Kumar, Kin On Kwok, Claudio F Lanata, Olivier Le Polain de Waroux, Kathy Leung, Wiriya Mahikul, Alessia Melegaro, Carl D Morrow, Joël Mossong, Eleanor FG Neal, David J Nokes, Wirichada Pan-ngum, Gail E Potter, Fiona M Russell, Siddhartha Saha, Jonathan D Sugimoto, Wan In Wei, Robin R Wood, Joseph T Wu, Juanjuan Zhang, Patrick GT Walker, Charles Whittaker

## Abstract

**Background:** Transmission of respiratory pathogens such as SARS-CoV-2 depends on patterns of contact and mixing across populations. Understanding this is crucial to predict pathogen spread and the effectiveness of control efforts. Most analyses of contact patterns to date have focussed on high-income settings.

**Methods:** Here, we conduct a systematic review and individual-participant meta-analysis of surveys carried out in low- and middle-income countries and compare patterns of contact in these settings to surveys previously carried out in high-income countries. Using individual-level data from 28,503 participants and 413,069 contacts across 27 surveys we explored how contact characteristics (number, location, duration and whether physical) vary across income settings.

**Results:** Contact rates declined with age in high- and upper-middle-income settings, but not in low-income settings, where adults aged 65+ made similar numbers of contacts as younger individuals and mixed with all age-groups. Across all settings, increasing household size was a key determinant of contact frequency and characteristics, but low-income settings were characterised by the largest, most intergenerational households. A higher proportion of contacts were made at home in low-income settings, and work/school contacts were more frequent in high-income strata. We also observed contrasting effects of gender across income-strata on the frequency, duration and type of contacts individuals made.

**Conclusions:** These differences in contact patterns between settings have material consequences for both spread of respiratory pathogens, as well as the effectiveness of different non-pharmaceutical interventions.

**Funding:** This work is primarily being funded by joint Centre funding from the UK Medical Research Council and DFID (MR/R015600/1).

## Introduction

Previous outbreaks of Ebola(Mbala-Kingebeni et al., 2019), influenza(Khan et al., 2009), and the ongoing COVID-19 pandemic have highlighted the importance of understanding the transmission dynamics and spread of infectious diseases, which depend fundamentally on the underlying patterns of social contact between individuals. Together, these patterns give rise to complex social networks that influence disease dynamics(Eubank et al., 2004; Ferrari et al., 2006; Firth et al., 2020; Zhang et al., 2020), including the capacity for emergent pathogens to become endemic(Ghani and Aral, 2005; Jacquez et al., 1988), the overdispersion of the offspring distribution underlying the reproduction number(Delamater et al., 2019) and the threshold at which herd-immunity is reached(Fontanet and Cauchemez, 2020; Mistry et al., 2021). They can similarly modulate the effectiveness of non-pharmaceutical interventions (NPIs), such as school closures and workplace restrictions, that are typically deployed to control and contain the spread of infectious diseases (Prem et al., 2020).

Social contact surveys provide insight into the features of these networks, which is typically achieved through incorporating survey results into mathematical models of infectious disease transmission frequently used to guide decision making in response to outbreaks(Chang et al., 2021; Davies et al., 2020). Such inputs are necessary for models to have sufficient realism to evaluate relevant policy questions. However, despite the known importance of contact patterns as determinants of the infectious disease dynamics, our understanding of how they vary globally remains far from complete. Reviews of contact patterns to date have focussed on High-Income countries (HICs)(Hoang et al., 2019). This is despite evidence that social contact patterns differ systematically across settings in ways that have material consequences for the dynamics of infectious disease transmission and the evolution of epidemic trajectories(Prem et al., 2017; Walker et al., 2020). Previous reviews has also primarily explored the total number of contacts made by individuals(Hoang et al., 2019) and/or how these contacts are distributed across different age/sex groups(Horton et al., 2020). Whilst these factors are a vital component underpinning disease spread, recent work has also underscored the importance of the characteristics of contacts (such as the location, duration and extent of physical contact) in determining transmission risk(Thompson et al., 2021).

Here, we carry out a systematic review of contact surveys (conducted prior to the emergence of COVID-19) in Lower-Income, Lower-Middle and Upper-Middle-Income countries (LICs, LMICs and UMICs, respectively). Alongside previously published data from HICs(Kwok et al., 2018, 2014; Leung et al., 2017; Mossong et al., 2008), we collate individual participant data (IPD) on social contacts from published work spanning 27 surveys from 22 countries and over 28,000 individuals. We use a Bayesian framework to explore drivers and determinants of contact patterns across a wider range of settings and at a more granular scale than has previously been possible. Specifically, we assess the influence of key factors such as age, gender and household structure on both the total number and characteristics (such as duration, location and type) of contact made by an individual, and explore how the comparative importance of different factors varies across different settings. We additionally evaluate the extent and degree of assortativity in contact patterns between different groups, and how this varies across settings.

## Results

### Systematic Review and Individual-Participant-Data (IPD) Meta-analysis

A total of 3,409 titles and abstracts were retrieved from the databases, and 313 full-text articles were screened for eligibility (Supplementary Figure 1). This search identified 19 studies with suitable contact data from LIC, LMIC and UMIC settings– individual-level data were obtained from 16 of these studies, including one study from a LIC, six studies from a LMIC and nine studies from an UMIC. These were analysed alongside four HIC studies from Hong Kong and Europe. Details of the identified studies and a full description of the systematic review findings can be found in Supplementary Text 1 and Supplementary Table 1. In total, this yielded 28,503 participants reporting on 413,069 contacts. All studies contained information on main demographic variables such as age and gender. Availability of other variables analysed here for each study are listed in Supplementary Table 2. All studies reported the number of contacts made in the past 24 hours of (or day preceding) the survey. The definitions of contacts were broadly similar across studies (Supplementary Table 1). Specifically, contacts were defined as skin-to-skin (physical) contact or a two-way conversation in the physical presence of another person. All studies scored above 65% of the items on the AXIS risk of bias tool, suggesting good or fair quality (Supplementary Table 3). Among all participants 47.5% were male, 30.1% were aged under 15 years and 7.2% were aged over 65 years. The majority (83.4%) of participants were asked to report the number of contacts they made on a weekday. A large proportion (34.1%) of respondents lived in large households of 6 or more people but this was largely dependent on income setting (LIC/LMIC=63.2%, UMIC=35.9%, HIC=4.9%). Among school-aged children (5 to 18 years), 88.1% were students, and 59.1% of adults aged over 18 were employed.

### Total number of contacts and contact location

The median number of contacts made per day across all the studies was 9 (IQR= 5-17), and was similar across income strata (LIC/LMIC=10[5-17], UMIC=8[5-16], HIC=9[5-17]; Table 1). There was a large variation in contact rates across different studies, with the median number of daily contacts ranging from 4 in a Zambian setting(Dodd et al., 2015) to 24 in an online Thai survey(Stein et al., 2014). When stratifying by study methodology, median daily contacts was higher in diary-based surveys compared to interview-/questionnaire-based surveys, which was true across all income strata (Table 1, Supplementary Figure 2).

**Table 1.** Summary table of total daily contacts. The total number of observations, as well as the mean, median and interquartile range (p25 and p75) of total daily contacts shown by participant and study characteristics.

|  |  |  | N | Mean | p25 | Median | p75 |
| --- | --- | --- | --- | --- | --- | --- | --- |
| **Overall** |  |  | 28,503 | 14.5 | 5 | 9 | 17 |
| **Gender** |  | **Male** | 13,218 | 15.3 | 5 | 9 | 18 |
|  |  | **Female** | 14,598 | 13.7 | 5 | 9 | 16 |
| **Age** |  | **<15** | 8,561 | 14.6 | 6 | 10 | 19 |
|  |  | **15 to 65** | 17,841 | 14.9 | 5 | 9 | 17 |
|  |  | **>65** | 2,047 | 10.4 | 3 | 6 | 12 |
| **Income status** |  | **LIC/LMIC** | 9,906 | 15.4 | 5 | 10 | 17 |
|  |  | **UMIC** | 8,330 | 14.4 | 5 | 8 | 16 |
|  |  | **HIC** | 10,267 | 13.7 | 5 | 9 | 17 |
| **Survey Methodology** |  | **Diary** | 12,226 | 13.9 | 6 | 10 | 18 |
|  |  | **Interview/Survey** | 16,227 | 15.0 | 4 | 8 | 16 |
| **Day type** |  | **Weekend** | 4,308 | 14.7 | 5 | 9 | 16 |
|  |  | **Weekday** | 21,579 | 14.1 | 5 | 9 | 17 |
| **Employment**<br>*(in those aged >18)* |  | **Yes** | 8,879 | 15.4 | 5 | 9 | 17 |
|  |  | **No** | 6,158 | 9.8 | 4 | 7 | 12 |
| **Student**<br>*(in those aged 5 to 18)* |  | **Yes** | 4,438 | 18.4 | 8 | 14 | 24 |
|  |  | **No** | 600 | 10.4 | 5 | 8 | 14 |
| **Household size** |  | **1** | 1,479 | 10.4 | 3 | 6 | 12 |
|  |  | **2** | 3,220 | 11.8 | 4 | 7 | 14 |
|  |  | **3** | 4,130 | 12.0 | 4 | 7 | 14 |
|  |  | **4** | 5,240 | 13.4 | 5 | 8 | 17 |
|  |  | **5** | 3,109 | 12.5 | 4 | 8 | 14 |
|  |  | **6+** | 8,873 | 17.7 | 7 | 11 | 20 |
| **Study** | **Belgium** | **Mossong** | 750 | 11.8 | 5 | 9 | 15 |
|  | **China** | **Read** | 1,821 | 18.6 | 7 | 13 | 22 |
|  | **China** | **Zhang** | 965 | 18.8 | 4 | 10 | 30 |
|  | **Fiji** | **Neal** | 2,019 | 6.4 | 4 | 6 | 8 |
|  | **Finland** | **Mossong** | 1,006 | 11.1 | 5 | 9 | 15 |
|  | **Germany** | **Mossong** | 1,341 | 7.9 | 4 | 6 | 10 |
|  | **Hong Kong** | **Kwok (2014)** | 762 | 18.3 | 5 | 9 | 18 |
|  | **Hong Kong** | **Kwok (2018)** | 1,066 | 11.9 | 3 | 7 | 13 |
|  | **Hong Kong** | **Leung** | 1,149 | 14.4 | 3 | 7 | 15 |
|  | **India** | **Kumar** | 2,943 | 27.0 | 12 | 17 | 26 |
|  | **Italy** | **Mossong** | 849 | 19.8 | 10 | 17 | 27 |
|  | **Kenya** | **Kiti** | 568 | 17.7 | 10 | 15 | 23 |
|  | **Luxembourg** | **Mossong** | 1,051 | 17.5 | 8 | 14 | 24 |
|  | **Netherlands** | **Mossong** | 269 | 13.9 | 6 | 11 | 19 |
|  | **Peru** | **Grijalva** | 588 | 15.3 | 8 | 12 | 20 |
|  | **Poland** | **Mossong** | 1,012 | 16.3 | 7 | 13 | 22.5 |
|  | **Russia** | **Ajelli** | 502 | 18.0 | 6 | 11 | 19 |
|  | **South Africa** | **Dodd** | 1,276 | 5.2 | 4 | 5 | 7 |
|  | **South Africa** | **Wood** | 571 | 15.6 | 9 | 14 | 20 |
|  | **Senegal** | **Potter** | 1,417 | 19.7 | 10 | 15 | 25 |
|  | **Thailand** | **Mahikul** | 369 | 22.6 | 13 | 20 | 31 |
|  | **Thailand** | **Stein** | 219 | 58.5 | 15 | 24 | 55 |
|  | **Uganda** | **Le Polain de Waroux** | 568 | 7.0 | 5 | 7 | 9 |
|  | **United** | **Mossong** | 1,012 | 11.7 | 6 | 10 | 16 |
|  | **Vietnam** | **Horby** | 865 | 7.7 | 5 | 7 | 9 |
|  | **Zambia** | **Dodd** | 2,300 | 4.8 | 3 | 4 | 6 |
|  | **Zimbabwe** | **Melegaro** | 1,245 | 10.7 | 6 | 9 | 14 |

Overall, children aged 5 to 15 had the highest number of daily contacts (Figure 1A-C), although there was substantial variation between studies and across income-strata in how the number of daily contacts varied with age (Figure 1A-C). Across UMICs and HICs, the number of daily contacts made by participants decreased with age, with this decrease most notable in the oldest age-groups (adjCRR for 65+ vs. <15 years [95%CrI]: UMIC=0.67[0.63-0.71] and HIC=0.57[0.54-0.60]). By contrast, there was no evidence of contact rates declining in the oldest age-groups in LICs/LMICs (adjCRR for 65+ vs. <15 years [95%CrI]=0.94[0.89-1.00]). We observed contrasting effects of gender on the number of daily contacts, with men making more daily contacts compared to women in LICs/LMICs after accounting for age (adjCRR=1.17, 95%CrI:1.15-1.20; Figure 1D), but no effect of gender on total daily contacts for other income strata (CRR[95%CrI]: UMIC=1.01[0.98-1.04], HIC=0.99[0.97-1.02]). There were also differences in the number of daily contacts made according to the methodology used and whether the survey was carried out on a weekday or over the weekend – in both instances, contrasting effects of these factors on the number of daily contacts according to income strata were observed (Figures 1D-1F).

**Figure 1.**
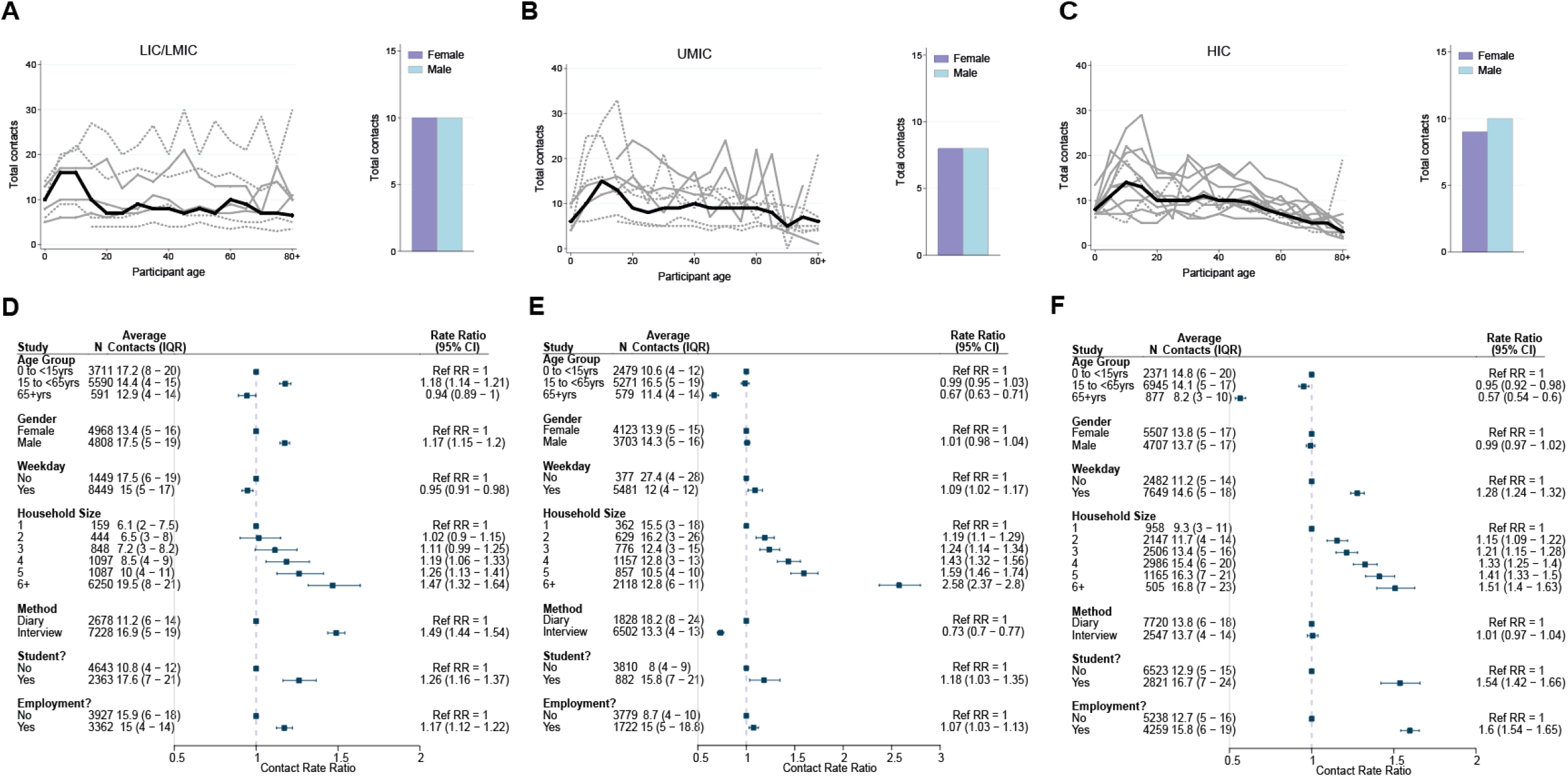
Total number of contacts. Sample median total number of contacts shown by gender (right) and 5-year age groups up to ages 80+ shown for A) LICs/LMICs, B) UMICs and C) HICs. Grey lines denote individual studies, and the solid black line is the median across all studies of within that income group. Studies with a diary-based methodology are represented by a solid grey line and those with a questionnaire or interview design are shown as a dashed line. For UMICs, one study outlier with extremely high number of contacts is excluded (online Thai survey with a “snowball” design by Stein et al., 2014). Contact Rate Ratios and associated 95% Credible intervals from a negative binomial model with random study effects are shown in D (LICs/LMICs), E (UMICs) and F (HICs).

We also examined the influence of factors that might influence both the total number and location (home, work, school and other) of the contacts individuals make. Across all income-strata, students (defined as those currently in education, attending school and aged between 5 and 18 years) made more daily contacts than non-students aged between 5 and 18 (adjCRR [95%CrI]: LIC/LMIC=1.26[1.16-1.37], UMIC=1.18[1.03-1.35] and HIC=1.54[1.42-1.66]; Figure 1D-F). Similarly, we observed strong and significant effects of employment in all income strata, with adults who were employed having a higher number of total daily contacts compared to those not in employment (adjCRR [95%CrI]: LIC/LMIC= 1.17[1.12-1.23], UMIC= 1.07[1.03-1.13], HIC= 1.60[1.54-1.65]; Figure 1D-F). Total daily contacts increased with household size (Figure 2A, Supplementary Figure 2) across all income-strata; individuals living in large households (6+ members) had 1.47 (95%CrI:1.32-1.64) (LIC/LMICs), 2.58 (95%CrI:2.37-2.80) (UMICs) and 1.51 (95%CrI:1.40-1.63) (HICs) times more daily contacts than those living alone, after accounting for age and gender (Figure 1E-F). Sensitivity analyses excluding additional contacts (as defined in Methods), showed little difference in effect sizes, and were strongly correlated with the effect sizes shown in Figure 1D-F (Supplementary Figure 3).

**Figure 2.**
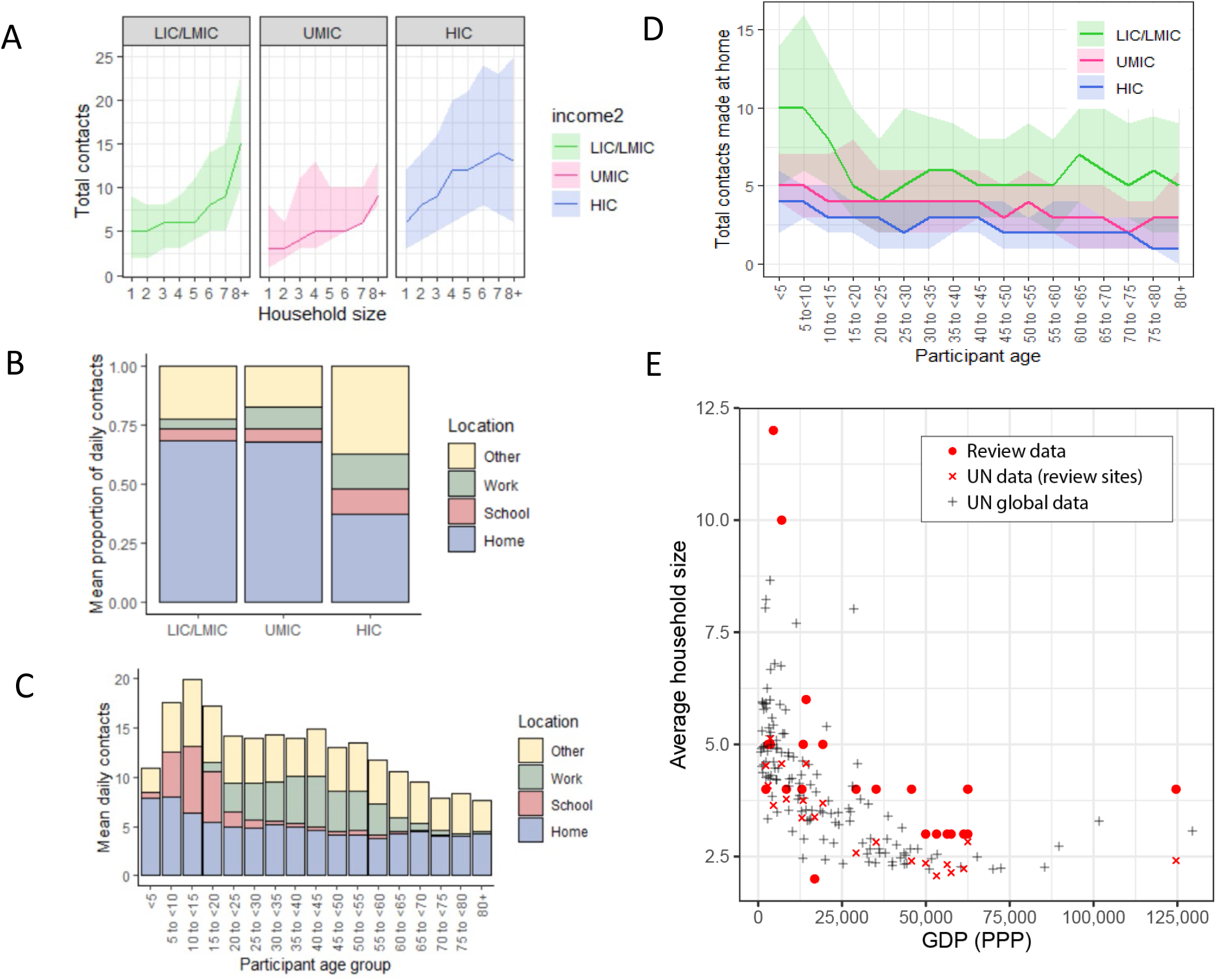
A) Sample median number of contacts by household size in review data, stratified by income strata. Shaded area denotes the interquartile range. B) sample mean % of contacts made at each location (home, school, work, other) by income group. C) total daily contacts (sample mean number) made at each location by 5-year age group. D) Sample median number of contacts made at home by 5-year age groups and income strata. Shaded area denotes the interquartile range. E) Average household size and GDP; red circles represent median household size in single studies from the review. GDP information was obtained from the World Bank Group and global household size data from the Department of Economic and Social Affairs, Population Division, United Nations.

Motivated by this suggestion of strong, location-related (school, work and household) effects on total daily contact rates, we further explored the locations in which contacts were made. Contact location was known for 314,235 contacts, 42.7% of which occurred at home (13.1% at work, 12.5% at school and 31.7% in other locations). Across income-strata, there was significant variation in the proportion of contacts made at home – being highest in LICs/LMICs (68.3%) and lowest in HICs (37.0%) (Figure 2B). Age differences were also observed in the number of contacts made at home, particularly for LICs/LMICs (Figure 2C-2D). Relatedly, a higher proportion of contacts occurred at work and school (14.6 % and 11.3%) in HICs compared to LICs/LMICs (3.9% and 5.2%, respectively; Supplementary Figure 4). Strong, gender specific patterns of contact location were also observed. Across all income strata males made a higher proportion of their contacts at work compared to females, although this difference was largest for LICs/LMICs (Supplementary Figure 4). Further, we found significant variation between income strata in median household size (7 in LICs/LMICs, 5 in UMICs and 3 in HICs). This trend of decreasing household size with increasing country income was consistent with global data (Figure 2E). The larger households observed for LIC/LMIC settings were also more likely to be intergenerational – in LICs/LMICs, 59.4% of participants aged over 65 lived in households of at least 6 members compared to 17.5% in UMICs and only 2.2% in HICs.

### Type and duration of contact

Data on the type of contacts (physical and non-physical) were recorded for 20,910 participants. The mean percentage of physical contacts across participants was 56.0% and was the highest for LICs/LMICs (64.5%). At the study level, the highest mean percentage of physical contacts was observed for a survey of young children and their caregivers conducted in Fiji(Neal et al., 2020) (84.0%) and the lowest in a Hong Kong contact survey(Leung et al., 2017)(18.9%). Physical contact was significantly less common among adults compared to children under 15 years in all settings (ORs ranged between 0.22 to 0.48) (Figure 3A-F). Despite the proportion of physical contacts generally decreasing with age, there was a higher proportion observed for adults aged 80 or over (Figure 3A-C). Contacts made by male participants were more likely to be physical compared to female participants in UMICs (adjOR= 1.13, 95%CrI=1.10-1.16) and HICs (adjOR= 1.09, 95%CrI=1.07-1.12), but in LICs/LMICs men had a lower proportion of physical contacts than women (adjOR= 0.81, 95%CrI=0.79-0.83; Figure 3D-F). Most physical contacts made by women in LICs were made at home (73.5%), whilst for HICs this was just 41.4% - similar differences across income-strata were observed for men, although the proportions were always lower than observed for women (62.4% for LIC/LMICs and 36.4% for HICs). Increasing household size was generally associated with a higher proportion of contacts being physical (for households of 6+ members compared to 1 member: adjCRR[95%CrI]: LIC/LMIC=1.73[1.48-2.02], UMIC= 1.30[1.12-1.52], HIC= 1.57[1.48-1.67]; Figure 3D-F). Employment was associated with having a significantly lower proportion of physical contacts in LICs/LMICs (adjOR=0.83, 95%CrI:0.79-0.87) and HICs (adjOR=0.71, 95%CrI:0.69-0.73), but not in UMICs (adjOR=1.11, 95%CrI:1.03-1.19). The proportion of physical contacts among all contacts was the highest for households (70.4%), followed by schools (58.5%), community (55.7%) and work (33.6%) (Supplementary Figure 5).

**Figure 3.**
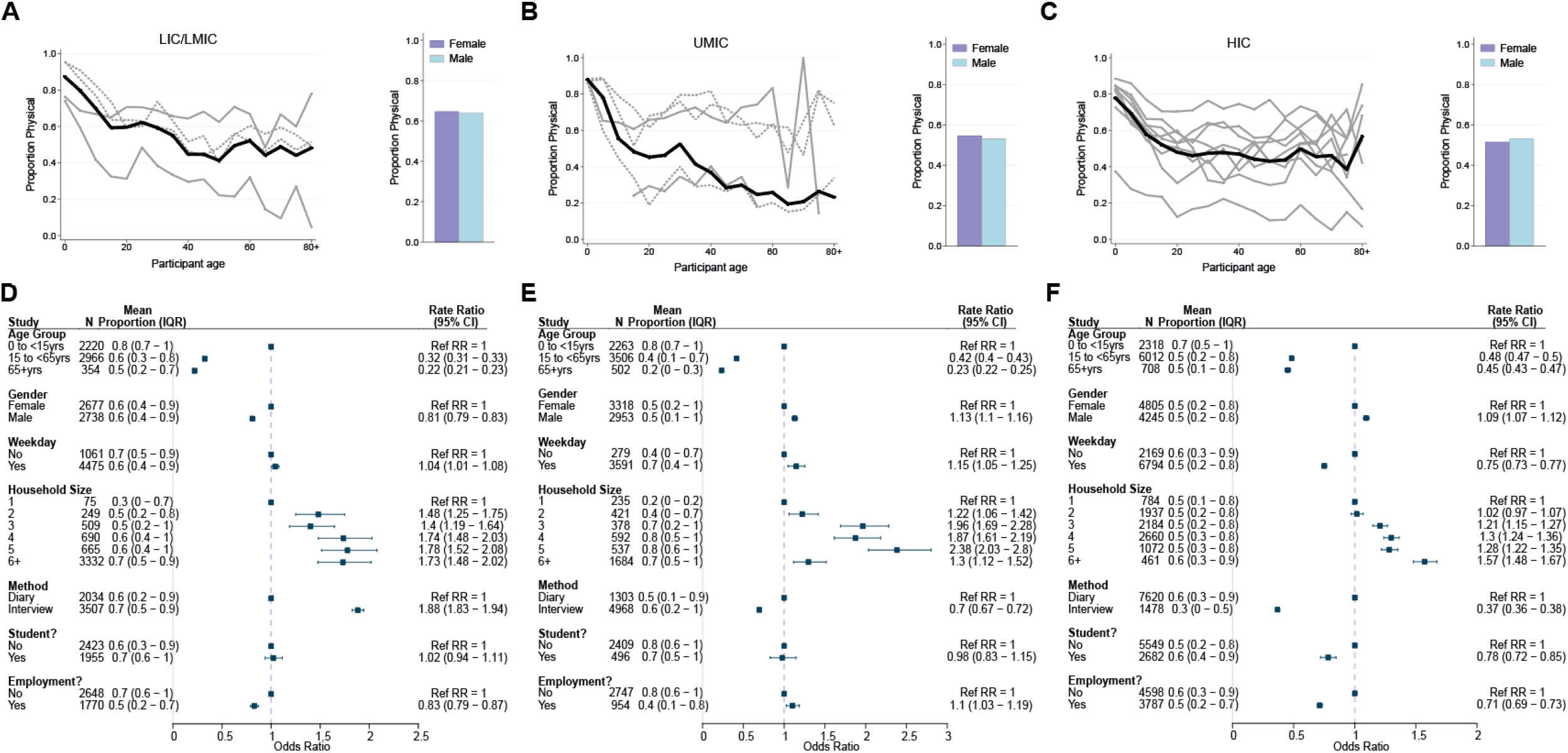
Physical contacts. Mean proportion of contacts that are physical shown by gender (right) and 5-year age groups up to ages 80+ shown for A) LICs/LMICs, B) UMICs and C) HICs. Grey lines denote individual studies, and the solid black line is the mean across all studies of within that income group. Studies with a diary-based methodology are represented by a solid grey line and those with a questionnaire or interview design are shown as a dashed line. Odds Ratios and associated 95% Credible intervals from a logistic regression model with random study effects are shown in D (LICs/LMICs), E (UMICs) and F (HICs).

Data on the duration of contact (<1 or ≥1hr) were available for 22,822 participants. The percentage of contacts lasting at least 1 hour was 63.2% and was highest for UMICs (76.0%) and lowest for LICs/LMICs (53.1%). Across both UMICs and HICs, duration of contacts was lower in individuals aged over 15 years compared to those aged 0-15, with the extent of this disparity most stark for HICs (for ages 65+ compared to <15 years: adjCRR [95%CrI]: LIC/LMIC= 0.61[0.57-0.64], UMIC= 0.61[0.58-0.65], HIC= 0.35[0.33-0.37]; Figure 4A-F). We observed contrasting effects of gender across income-strata: males made longer-lasting contacts than females in UMICs (adjOR=1.11, 95%CrI=1.08-1.14); Figure 4D-F), but not in LIC/LMICs (adjOR=0.92, 95%CrI=0.90-0.95) or HICs (adjOR=0.98, 95%CrI=0.97-1.00). Participants reported shorter contacts on weekends compared to weekdays in LICs/LMICs (adjOR=0.91, 95%CrI: 0.88-0.95), and HICs (adjOR=0.95, 95%CrI: 0.92-0.97), but not in UMICs (adjOR=1.12, 95%CrI=1.03-1.21). Contacts lasting over an hour as a proportion of all contacts was highest for households (72.7%), followed by schools (67.9%), community (47.0%) and work (44.0%). However, it was only in HICs that there was a significant effect of being a student (adjOR=1.18, 95%CrI: 1.09-1.27; Figure 4D-F) on the proportion of contacts lasting ≥1 hour. For all income strata, the proportion of contacts >1h increased with increasing household size (Figure 4D-F).

**Figure 4.**
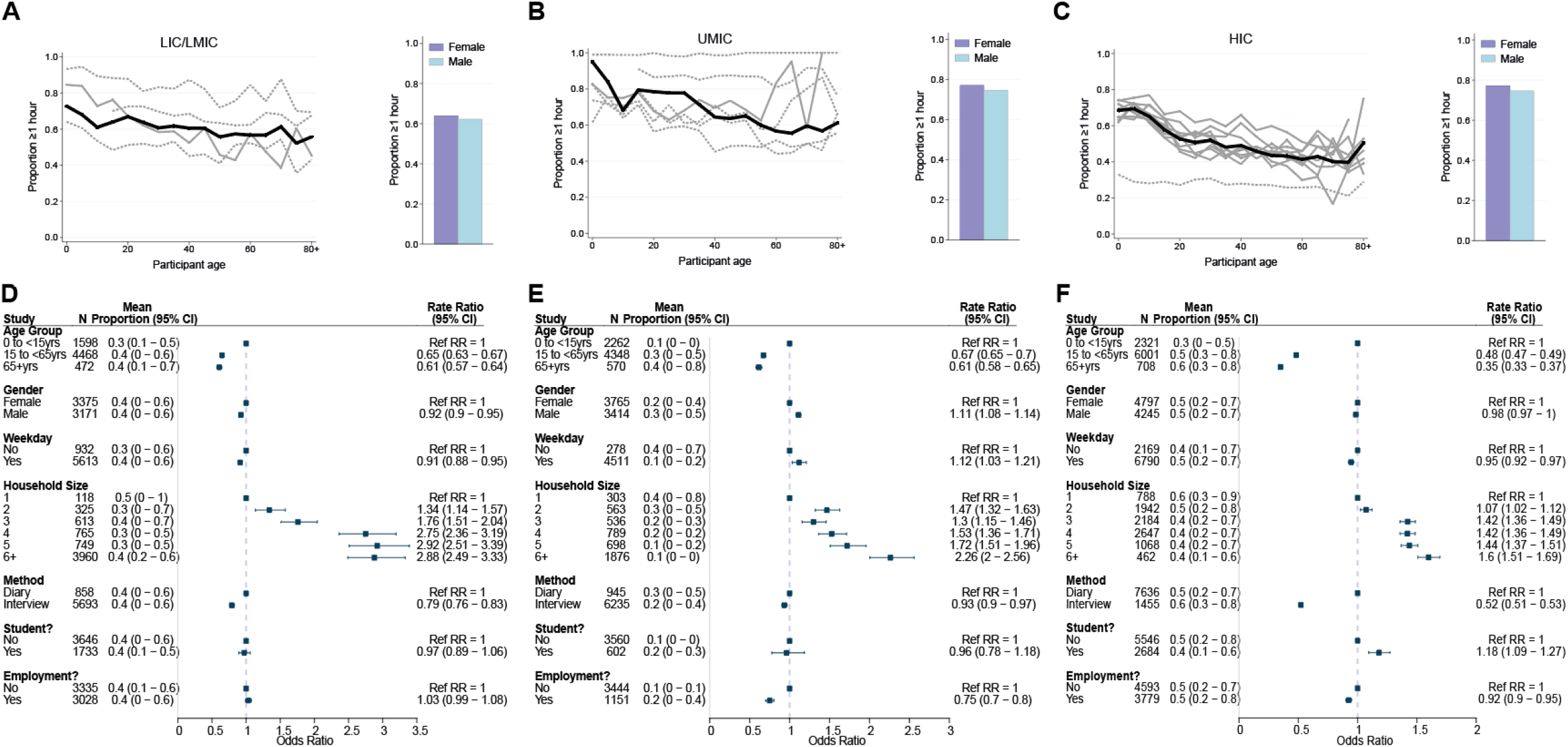
Contact duration. Mean proportion of contacts that last at least an hour shown by gender (right) and 5-year age groups up to ages 80+ shown for A) LICs/LMICs, B) UMICs and C) HICs. Grey lines denote individual studies and the solid black line is the mean across all studies of within that income group. Studies with a diary-based methodology are represented by a solid grey line and those with a questionnaire or interview design are shown as a dashed line. Odds Ratios and associated 95% Credible intervals from a logistic regression model with random study effects are shown in D (LICs/LMICs), E (UMICs) and F (HICs).

### Assortativity by age and gender

Twelve studies collected information on the gender of the contact and eight studies contained information on age allowing assignment of contacts to one of the three age-groups described in Methods (Supplementary Table 2, Supplementary Text 2). We found evidence to suggest that contacts were assortative by gender for all income strata, as participants were more likely to mix with their own gender (Supplementary Text 2). Mixing was also assortative by age, with participants more likely to contact individuals who belonged to the same age group this degree of age-assortativity was lowest for LICs/LMICs, where only 29% of contacts made by adults were with individuals of the same age group. By contrast, in HICs we observed a higher degree of assortative mixing, with most contacts (51.4%) made by older adults occurring with individuals belonging to the same age group.

## Discussion

Understanding patterns of contact across populations is vital to predicting the dynamics and spread of infectious diseases, as well understanding the control interventions likely to have the greatest impact. Here, using a systematic review and individual-participant data meta-analysis of contact surveys, we summarise research exploring these patterns across a range of populations spanning 28,503 individuals and 22 countries. Our findings highlight substantial differences in contact patterns between income settings. These differences are driven by setting-specific sociodemographic factors such as age, gender, household structure and patterns of employment, which all have material consequences for transmission and spread of respiratory pathogens.

Across the collated studies, the total number of contacts was highest for school-aged children. This is consistent with previous results from HICs(Béraud et al., 2015; Fu et al., 2012; Hoang et al., 2019; Ibuka et al., 2016; Lapidus et al., 2013) and shown here to be generally true for LICs/LMICs and UMICs also. Interestingly however, we observed differences in patterns of contact in adults across income strata. Whilst contact rates in HICs declined in older adults, this was not observed in LICs/LMICs, where contact rates did not differ in the oldest age-group compared to younger ages. This is consistent with variation in household structure and size across settings, with nearly two thirds of participants aged 65+ in included LIC/LMIC surveys living in large, likely intergenerational, households (6+ members), compared to only 2% in HICs. HICs were also characterised by more assortative mixing between age-groups, with older adults in LICs/LMICs more likely to mix with individuals of younger ages, again consistent with the observed differences between household structures across the two settings. These results have important consequences for the viability and efficacy of protective policies centred around shielding of elderly individuals (i.e. those most at risk from COVID-19 or influenza) in these settings.

Our results support the idea of households as a key site for transmission of respiratory pathogens(Thompson et al., 2021), with the majority of contacts made at home. However, its relative importance compared to other locations is likely to vary across settings. Our results highlighted significant differences across income settings in the distribution of contacts made at home, work and school. The proportion of contacts made at home was highest for LIC/LMICs, where larger average household sizes were associated with more contacts, more physical contacts, and longer lasting contacts. By contrast, participants in HICs tended to report more contacts occurring at work and school. The lower number of contacts at work in LIC/LMIC may be explained by the types of employment (e.g agriculture in rural surveys) and a selection bias (women at home/homemakers more likely to be surveyed in questionnaire-based surveys). Such differences would have consequences for which locations contribute most to transmission and in turn modulate the efficacy of different NPIs, such as workplace closures. Our analyses similarly highlighted significant variation in the duration and nature of contacts across settings. Contacts made by female participants in LICs/LMICs were more likely to be physical compared to men, whilst the opposite effect was observed for HICs and UMICs, potentially reflecting context-specific gender roles. In all settings, we observed a general decline of physical contacts with age, except in the very old(Mossong et al., 2008), potentially reflecting higher levels of dependency and the need for physical care.

There are important caveats to these findings. Data constraints limited the numbers of factors we were able to explore – for example, despite evidence(Kiti et al., 2014) suggesting that contact patterns differ across rural and urban settings, only 3 studies(Kiti et al., 2014; O. le Polain de Waroux et al., 2018; Neal et al., 2020) contained information from both rural and urban sites, allowing classification. Similarly, we were unable to examine the impact of socioeconomic factors such as household wealth, despite experiences with COVID-19 having highlighted strong socio-economic disparities in both transmission and burden of disease(De Negri et al., 2021; Routledge et al., 2021; Ward et al., 2021; Winskill et al., 2020) and previous work suggesting that poorer individuals are less likely to be employed in occupations amenable to remote working(Loayza, 2020). A lack of suitably detailed information in the studies conducted precludes analysis of these factors but highlights the importance of incorporating economic questions into future contact surveys, such as household wealth and house square footage. Other factors also not controlled for here, but that may similarly shape contact patterns include school holidays or seasonal variations in population movement and composition that we are unable to capture given the cross-sectional nature of these studies.

Another important limitation to the results presented here is that we are only able to consider a limited set of contact characteristics (the location and duration of the contact and whether it was physical). Previous work has highlighted the importance of these factors in determining the risk of respiratory pathogen transmission(Chang et al., 2021; Dunne et al., 2018; Olivier le Polain de Waroux et al., 2018; Neal et al., 2020; Thompson et al., 2021), but only a limited number of studies reported whether a contact was “close” or “casual”(Kwok et al., 2018, 2014; O. le Polain de Waroux et al., 2018) and whether the contact was made indoors or outdoors(Wood et al., 2012); both factors likely to influence transmission risk(Bulfone et al., 2021; Chu et al., 2020). More generally, the relevance and comparative importance of different contacts to transmission likely varies according to the specific pathogen and its predominant transmission modality (e.g. aerosol, droplet, fomite etc). It is therefore important to note that these results do not provide a direct indication of explicit transmission risk, but rather an indicator of factors likely to be relevant to transmission. Relatedly, it is also important to note that the studies collated here were all conducted prior to the onset of the SARS-CoV-2 pandemic. Previous work has documented significant alterations to patterns of social contact in response to individual-level behaviour changes or government implemented NPIs aimed at controlling SARS-CoV-2 spread, but detailed analysis of changing contact patterns is contingent on both an understanding of baseline contact patterns as detailed in the studies collated here as well as longitudinal sampling of how contacts patterns change over time, which is available for only a limited number of settings(Jarvis et al., 2021, 2020; Liu et al., 2021). Description of contact location was also coarse and precluded more granular analyses of specific settings, such as markets, which have previously been shown to be important locations for transmission in rural areas(Grijalva et al., 2015).

Heterogeneity between studies was larger for LICs/LMICs and UMICs, which we partly accounted for, through fitting random study effects. These study differences may be attributed to the way individual contact surveys were conducted, making comparisons of contact patterns among surveys more difficult (e.g. prospective/retrospective diary surveys, online/paper questionnaires, face-to-face/phone interviews, and different contact definitions). For instance, there is evidence suggesting that prospective reporting, which is less affected by recall bias, can often lead to a higher number of contacts being reported(Mikolajczyk and Kretzschmar, 2008) and a lower probability of casual or short-lasting contacts being missed. The relatively high contact rates observed in HICs may be explained by the fact that all but two HIC surveys used diary methods. Our study highlights that a unified definition of “contact” and standard practice in data collection could help increase the quality of collected data, leading to more robust and reliable conclusions about contact patterns. Whilst we aggregate results by income strata due to the limited availability of data (particularly in lower- and middle-income countries), it is important to note that the outcomes considered here are likely to be shaped by several different factors other than country-level income. Whilst some of these factors will be correlated with a country’s income status (e.g. household size(Walker et al., 2020)), many others however will be unique to a particular setting or geographical area or correlate only weakly with country-level data. Examples include patterns of employment, the role of women, and other contextual factors. These analyses are therefore intended primarily to provide indications of prevailing patterns, rather than a definitive description of contact patterns in a specific context and highlight the significant need for further studies to by carried out in a diversity of different locations.

Despite these limitations however, our results highlight significant differences in the structure and nature of contact patterns across settings. These differences suggest that the comparative importance of different locations and age-groups to transmission will likely vary across settings and have critical consequences for the efficacy and suitability of strategies aimed at controlling the spread of respiratory pathogens such as SARS-CoV-2. Most importantly, our study highlights the limited amount of work that has been undertaken to date to better understand and quantify patterns of contact across a range of settings, particularly in lower- and middle-income countries, which is vital in informing control strategies reducing the spread of such pathogens.

## Methods

### Systematic Review

#### Data sources and search strategy

Two databases (Ovid MEDLINE and Embase) were searched on 26^th^ May 2020 to identify studies reporting on contact patterns in LICs, LMICs and UMICs (Supplementary Table 4). Collated records underwent title and abstract screening for relevance, before full-text screening using pre-determined criteria. Studies were included if they reported on any type of face-to-face or close contact with humans and were carried out in LICs, LMICs or UMICs only. No restrictions on collection method (e.g. prospective diary-based surveys or retrospective surveys based on a face-to-face/phone interview or questionnaire) were applied. Studies were excluded if they did not report contacts relevant to air-borne diseases (e.g. sexual contacts), were conducted in HICs, were contact tracing studies of infected cases, or were conference abstracts. All studies were screened independently by two reviewers (AM and CW). Differences were resolved through consensus and discussion. The study protocol can be accessed through PROSPERO (registration number: CRD42020191197). Income group classification (LIC/LMIC, UMIC, or HIC) was based on 2019 World Bank data (fiscal year 2021)(World Bank Group, 2020).

#### Data extraction

Individual-level data were obtained from publication supplementary data, as well as online data repositories such as Zenodo, figshare and OSF. When not publicly available, study authors were contacted to request data. Extracted data included the participant’s age, gender, employment, student status, household size and total number of contacts, as well as the day of the week for which contacts were reported. Some studies reported information at the level of individual contacts and included the age, gender, location and duration of the contact, as well whether it involved physical contact. Individual-level data from HICs, not systematically identified, were used for comparison, and included three studies from Hong Kong(Kwok et al., 2018, 2014; Leung et al., 2017) and the 8 European countries from the POLYMOD study(Mossong et al., 2008). Data were collated, cleaned and standardised using Stata version 14. Country-specific average household size were obtained from the United Nations Database on Household Size and Composition(United Nations Department of Economic and Social Affairs Population Division, 2019). Gross domestic product based on purchasing power parity (GDP PPP) was obtained from the World Data Bank database(World Bank International Comparison Programme, 2021). Findings are reported in accordance with the Preferred Reporting Items for Systematic Reviews and Meta-Analyses (PRISMA) checklist of items specific to IPD meta-analyses (Supplementary Table 5). Risk of bias was assessed using the AXIS critical appraisal tool used to evaluate quality of cross-sectional studies(Downes et al., 2016), modified to this study’s objectives (Supplementary Table 3). Each item was attributed a zero or a one, and a quality score was assigned to each study, ranging from 0% (“poor” quality) to 100% (“good” quality). The individual-level data across all studies and analysis code are available at https://github.com/mrc-ide/contact_patterns (see Supplementary Text 3 for data dictionary).

### Statistical analysis

The mean, median and interquartile range of total daily unique contacts were calculated for subgroups including country income status, individual study, survey methodology (diary-based or questionnaire/interview-based), survey day (weekday/weekend), and respondent characteristics such as age, sex, employment/student status and household size. Detailed description of data assumptions for each study can be found in Supplementary Text 3.

A negative binomial regression model was used to explore the association between the total number of daily contacts and the participant’s age, sex, employment/student status and household size, as well as methodology and survey day. Incidence rate ratios from these regressions are referred to as “Contact Rate Ratios” (CRRs). A sensitivity analysis was carried out that excluded additional contacts (such as additional work contacts, group contacts, and number missed out, which were recorded separately and in less detail by participants compared to their other contacts(Ajelli and Litvinova, 2017; Kumar et al., 2018; Leung et al., 2017; Zhang et al., 2020)). Logistic regressions were used to explore determinants of contact duration (<1hr/1hr+) and type (physical/non-physical), using the same explanatory variables as in the total contacts analyses. The proportion of contacts made at each location (home, school, work and other) was explored descriptively and contacts made with the same individual in separate locations/instances were considered as separate contacts.

All analyses were done in a Bayesian framework using the probabilistic programming language Stan, using uninformative priors in all analyses and implemented in R via the package *brm*s(Bürkner, 2018, 2017). All analyses were stratified by three income strata (LICs and LMICs were combined to preserve statistical power) and included random-study effects, apart from models adjusting for methodology which did not vary by study. The effect of each factor was explored in an age- and gender-adjusted model. All models exploring the effect of student status or employment status were restricted to children aged between 5 and 18 years and adults over 18, respectively. In the remaining models including all ages, age was adjusted as a categorical variable (<15, 15 to 65 and over 65 years). CRRs, Odds Ratios (ORs) and their associated 95% Credible Intervals are presented for all regression models. Here, we report estimates adjusted for age and gender (referred to as adjCRR or adjOR). Studies which collated contact-level data were used to assess assortativity of mixing by age and gender for different country-income strata by calculating the proportions of contacts made by participants that are male or female and those that belong to three broad age groups (children, adults, and older adults; Supplementary Text 2).

### Ethics statement

All original studies included were approved by an institutional ethics review committee. Ethics approval was not required for the present study.

## Data Availability

All data generated or analysed during this study are included in the manuscript and supporting files. Individual-level data across all studies and analysis code are available at https://github.com/mrc-ide/contact_patterns (see Supplementary Text 3 for data dictionary).

## Acknowledgements

We would like to acknowledge the Fiji Ministry of Health and Medical Services for their contribution to the study set in Fiji, M. Elizabeth Halloran for sharing the Senegal data, and Nickson Murunga for processing the data request for the Kenyan survey.

